# Heterogeneity of effect of Intensive lifestyle intervention on cardiometabolic risk factors by sex hormones in diabetes

**DOI:** 10.1101/2025.09.02.25334965

**Authors:** Chigolum P. Oyeka, Jianqiao Ma, Jiahuan Helen He, Nityasree Srialluri, Teresa Gisinger, Erin D. Michos, Mark Woodward, Rita R. Kalyani, Jeanne M. Clark, Wendy L. Bennett, Dhananjay Vaidya

## Abstract

**Background:** Intensive lifestyle intervention(ILI) in type 2 diabetes(T2D) improves cardiometabolic risk factors. Sex differences in these responses may be driven by the sex hormones testosterone(T), estradiol(E2), and sex hormone binding globulin(SHBG). We evaluated whether baseline sex hormone levels modify ILI effects on cardiometabolic risk factors ie, heterogeneity of treatment effect(HTE), and whether these modifications differ by sex.

**Methods:** Study included 2,260 Look AHEAD participants(1,093 postmenopausal females; 1,167 males, mean age 60 years) randomized to ILI or diabetes support and education with sex hormone measurements. We used linear mixed-effects models, stratified by sex and adjusted for age, race, study site, medications and baseline weight, to examine the association between baseline T, E2, and SHBG with change in lipids, hemoglobin A1c(HbA1c), systolic and diastolic blood pressure(BP), weight, and waist circumference(WC) over an 8-year follow-up. HTE was assessed using a three way interaction term between baseline hormone levels, time and randomization arm. Permutation tests controlled for multiple comparisons.

**Results:** In males, lower baseline E2 and total T significantly augmented ILI-induced triglyceride reductions(p=0.019 and 0.008, respectively) throughout follow up. Higher SHBG enhanced LDL-C lowering in females(p=0.018) and HDL-C increases in males(p=0.043). Higher baseline total T predicted greater long-term weight(p=0.013 females; 0.003 males) and WC loss(p=0.061 females; 0.035 males), while fewer hormone interactions were observed for BP and HbA1c.

**Conclusions:** Males with greater endogenous total T achieved larger reductions in triglycerides, weight, and WC, whereas females with higher total T had greater HDL-C improvements and those with higher SHBG experienced more BP lowering. Hormone profiling may guide personalized lifestyle prescriptions to improve long term cardiometabolic benefits.

**Clinical Perspective:**

- **What Is New?** Baseline sex hormone levels especially estradiol in males and testosterone/SHBG in females, sex-specifically modify the cardiometabolic benefits of intensive lifestyle intervention in type 2 diabetes.
- **What Are the Clinical Implications?** Endogenous hormone profiles may help clinicians personalize diet and exercise programs and guide adjunctive therapies to maximize lipid, blood pressure, and adiposity outcomes in females and males with T2D.

## Introduction

In people living with type 2 diabetes (T2D), intensive behavioral lifestyle interventions that result in weight loss lead to significant improvements in cardiometabolic risk factors, including lipid profiles (triglycerides [TG], low-density-lipoprotein cholesterol [LDL-C], high-density-lipoprotein cholesterol [HDL-C]), glycemic control (HbA1c)), blood pressure (systolic [SBP] and diastolic [DBP]), and waist circumference (WC))^1,2^. Importantly, sex differences in response to cardiometabolic risk after behavioral weight loss interventions have been previously reported^3–8^. Females experience greater improvement in fasting glucose, TG, and HDL-C and less improvement in HbA1c and LDL-C^4,7,8^. Comparatively, males tend to experience greater reductions in visceral adiposity and blood pressure, and less improvements in TG and HDL-C^4,5,7^. Sex differences in cardiometabolic risk factors may be moderated by biological factors, including hormonal changes and differential fat distributions, highlighting the complexity of sex-specific metabolic adaptations^9,10^.

Sex hormones (testosterone [T], estrogen [E2]) and sex hormone-binding globulin (SHBG)) are known to modulate lipid profiles, glucose metabolism, and fat distribution, which differentially affect cardiometabolic risk in females and males^11,12^. Variations in these hormone levels is associated with different female-male metabolic profiles, with low T in males and unusually low or abnormally high E2 levels in females being linked to adverse lipid and glucose outcomes^13^. Moreover, SHBG is associated with metabolic regulation, where lower concentrations correlate with T2D risk and cardiometabolic events^14–16^.

The Look AHEAD (Action for Health in Diabetes) randomized controlled trial provided a unique opportunity to investigate the long-term impacts of an intensive lifestyle intervention (ILI) for weight loss on a range of cardiometabolic risk factors in individuals with T2D and overweight or obesity. The trial enrolled over 5,000 adults with overweight and obesity and T2D and compared the effect of an ILI, emphasizing calorie restriction and exercise, vs a diabetes support and education (DSE) control on cardiometabolic morbidity and mortality ^17^. In the Sex Hormone ancillary study to Look AHEAD, we previously measured sex hormones (T, E2, and SHBG) and assessed the role of the intervention on sex hormones and any sex differences among postmenopausal females and older males living with T2D ^18^.

In the current study, we examined whether baseline sex hormone levels modify the response to the ILI targeting cardiometabolic risk factors: TG, LDL-C, HDL-C, HbA1c, SBP, DBP, weight, and WC. We also assessed whether there are differences in these responses by sex. We hypothesized that distinct baseline sex hormone profiles would predict differential trajectories in these cardiometabolic risk factors in response to lifestyle modifications. Elucidating the interactions between the ILI and sex hormones would advance our understanding of sex-specific metabolic adaptations in cardiometabolic risk in people with T2D, as an important step to personalize treatment for cardiometabolic disease prevention.

## Methods

### Study Design and Population

The Look AHEAD trial randomized 5,145 overweight or obese adults with T2D to ILI vs DSE and followed them for a median of 9.6 years, including a formal post-trial extension that has provided up to 8 years of additional follow-up for key cardiometabolic outcomes^19^. The Sex Hormone Ancillary Study was nested within this parent trial^8,18^. The Johns Hopkins School of Medicine IRB approved the study, and the Look AHEAD steering committee granted ancillary study approval.

### Sample Selection

In the Sex Hormone Ancillary Study, we selected a random sample of postmenopausal females (n=1093) and older males (n=1167, total n =2260) from the 5145 females (n=3063) and males (n=2082) in the main Look AHEAD trial to assess sex hormone levels (T, E2, and SHBG) over time^18^. Sample selection and baseline characteristics have been previously described^18,20^. Plasma samples were taken at 3 time points (baseline, year 1, and year 4)^18^. Exclusion criteria were missing stored samples at baseline, year 1 or year 4, females receiving breast cancer treatment or taking exogenous hormones at baseline, females < 55 years of age at baseline and either pre-menopausal or who had hysterectomy without oophorectomy, and males on anti-androgen medications for prostate cancer or androgen replacement at baseline^18^.

### Data Variables

#### Intervention

Participants were randomly assigned to ILI vs. DSE. The ILI comprised calorie restriction, physical activity, and frequent individual and group counselling (weekly in year 1) tapering off over the years of follow up^21,22^. The DSE arm received three group health education sessions annually without specific diet or exercise goals^21,23^.

#### Intervention Effect Modifiers

Sex hormones T, E2, and SHBG were the exposures. Laboratory methods for the sex hormones assessed in this ancillary study has been previously reported in detail^18,20^.Briefly, total T and E2 were measured using highly sensitive assays (negative electron capture chemical ionization gas chromatography-mass spectrometry [GCMS]) to ensure enhanced detection of the low levels in older adults^18,20^. SHBG was measured using ELISA (RRID: AB_3255149, Cat# K151G9 K, Mesoscale Discovery, MD)^18,20^.

### Outcomes: Cardiometabolic Risk Factors

All cardiometabolic risk factors were measured at baseline and years 1, 2, 3, 4, 6 and 8.

#### Lipids (TG, HDL-C, LDL-C)

Fasting blood samples were collected for lipid measurements, including TG, and HDL-C^21,24^. Lipid concentrations were measured using standardized enzymatic assays in a central laboratory, ensuring consistency across study sites^21^. LDL-C levels were calculated using Friedewald formula, which is accurate for triglycerides <400 mg/dL^24^. For participants with triglycerides ≥400 mg/dL, we performed a sensitivity analysis by excluding them.

#### Blood pressure

SBP and DBP were assessed with participants seated and after a 5-minute rest period; automated sphygmomanometers were used to obtain multiple readings, which were then averaged to improve measurement accuracy^21^.

#### Body weight and WC

Weight was measured in kg annually with a calibrated digital scale^25^. WC was measured twice and averaged at each annual visit with a non-stretchable measuring tape between the highest point of the iliac crest and the lowest part of the rib cage^21,26^.

#### Glycemic measures

HbA1c was measured from fasting blood specimens using high-performance liquid chromatography methods, calibrated according to Diabetes Control and Complications Trial (DCCT) standards to ensure reliable assessment of glycemic control^21,27^.

#### Other Covariates

Age, race, and study site were all from baseline measurements. Medication use (statins, antihypertension drugs, and insulins) was self-reported for time varying covariates.

### Statistical Analysis

We examined whether baseline sex hormone levels modified the effect of ILI on cardiometabolic outcomes over time using linear mixed-effects models. TG was log-transformed to ensure nearly normal distribution. We logarithmically transformed the sex hormone variables and divided them by their sex-specific standard deviation to allow for comparisons of treatment effect between sex hormones. Models were stratified by sex and adjusted for age, race, study site, and baseline weight. Models with lipids as outcomes were adjusted for statin use, models using weight, WC, and HbA1c as outcomes were adjusted for insulin use. Models using outcomes of SBP and DBP were adjusted by antihypertensive medication use. We evaluated heterogeneity of treatment effects using a three-way interaction term. This assessed the joint effects of baseline log-transformed sex hormones (E2, total T, SHBG), intervention (ILI vs. DSE), and follow-up year (1, 2, 3, 4, 6, 8 as categorical variables).

Sex hormone levels were modeled as continuous predictors in the statistical models. To visualize the estimated treatment effects, we plotted the predicted treatment effect in the cardiometabolic risk factors over time for two representative baseline hormone levels, corresponding to the 25th and 75th percentiles of baseline E2, total T, and SHBG, separately for males and females, <u>with error bars representing ±1 standard error.</u> Additionally, based on linear mixed-effects models above, we estimated the adjusted levels of the cardiometabolic variables for ILI and DSE groups for each follow-up year.

Analyses included multiple null hypothesis tests of effect modification, where the proposed effect modifiers, the sex hormone variables, covaried. Thus, multiple testing corrections based on uncorrelated null hypothesis, such as the Bonferroni correction are overly conservative. Because effect modification is for randomization group, a variable uncorrelated with any regression covariate by design, we generated empirical null distributions of the multiply tested 3-way interactions using 4000 random permutations of the randomized group variable. Diagnostic plots determined that this number of permutations produced stable results. We determined that only 5% of iterations had <=9 null hypothesis tests with p<0.05; thus, for this study if >=9 null hypothesis tests were p<0.05, the overall study had a false positive rate of <= 0.05. The multiple testing procedure has been detailed in supplementary material (Methods supplement).

To assess sex differences in the heterogeneity of the treatment effect (HTE) by baseline hormone levels from the sex-stratified regression models, we calculated the differences of each of the standardized hormone by ILI by categorical time interaction terms between female and male models, as differences in normally distributed variates with the estimated means and standard deviations, standardized by the standard error of the difference. The sum of all these standardized difference terms was tested as a chi-square variate with the appropriate number of degrees of freedom. For this exploratory analysis p<=0.05 was used as the level of statistical significance.

All statistical analyses were performed using Stata 18.0, and results with p<0.05 already corrected for multiple comparisons were highlighted.

## Results

**Supplement Figure 1** shows the selection process for the analytic sample. **Table 1** shows the baseline sample characteristics of 1093 females (ILI: 551, DSE: 542) and 1167 males (ILI: 589, DSE: 578).. The mean age was 60 years. There were more Black females (vs. Black males) in both intervention arms (Black females: ILI, 18.5%, DSE, 22.5%, vs. Black males: ILI: 9.5%, DSE: 9.3%). Across the two arms, females had lower median TG (146.0 vs. 163.0 mg/dL) and higher mean HDL-C (46.7 vs. 38.1 mg/dL), but higher mean LDL-C (118.4 vs. 106.5 mg/dL) compared to males. Females had higher mean BMI (36.3 vs. 34.9 kg/m^2^) but lower WC (110.2 vs. 117.8 cm) compared to males. HbA1c was similar between sexes (mean 7.2%). SBP was comparable, while females had lower DBP (67.6 vs. 73.1 mmHg).

**Table 1.**
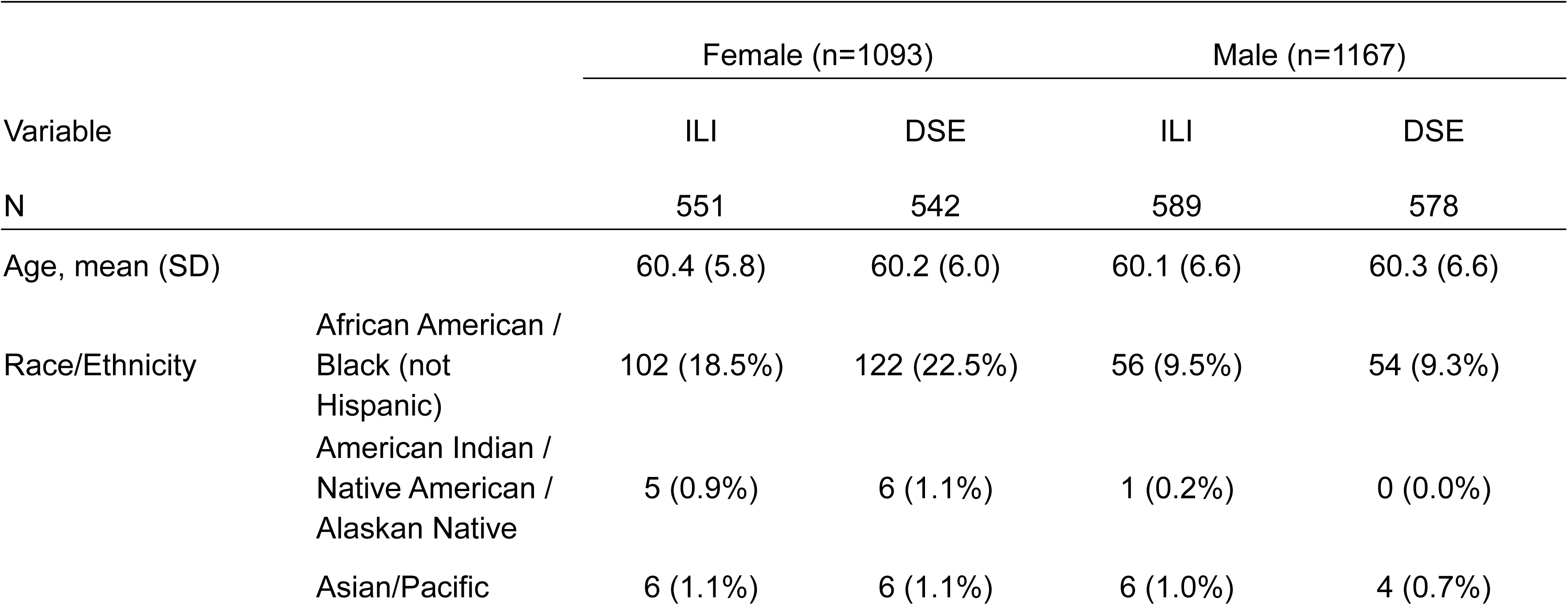
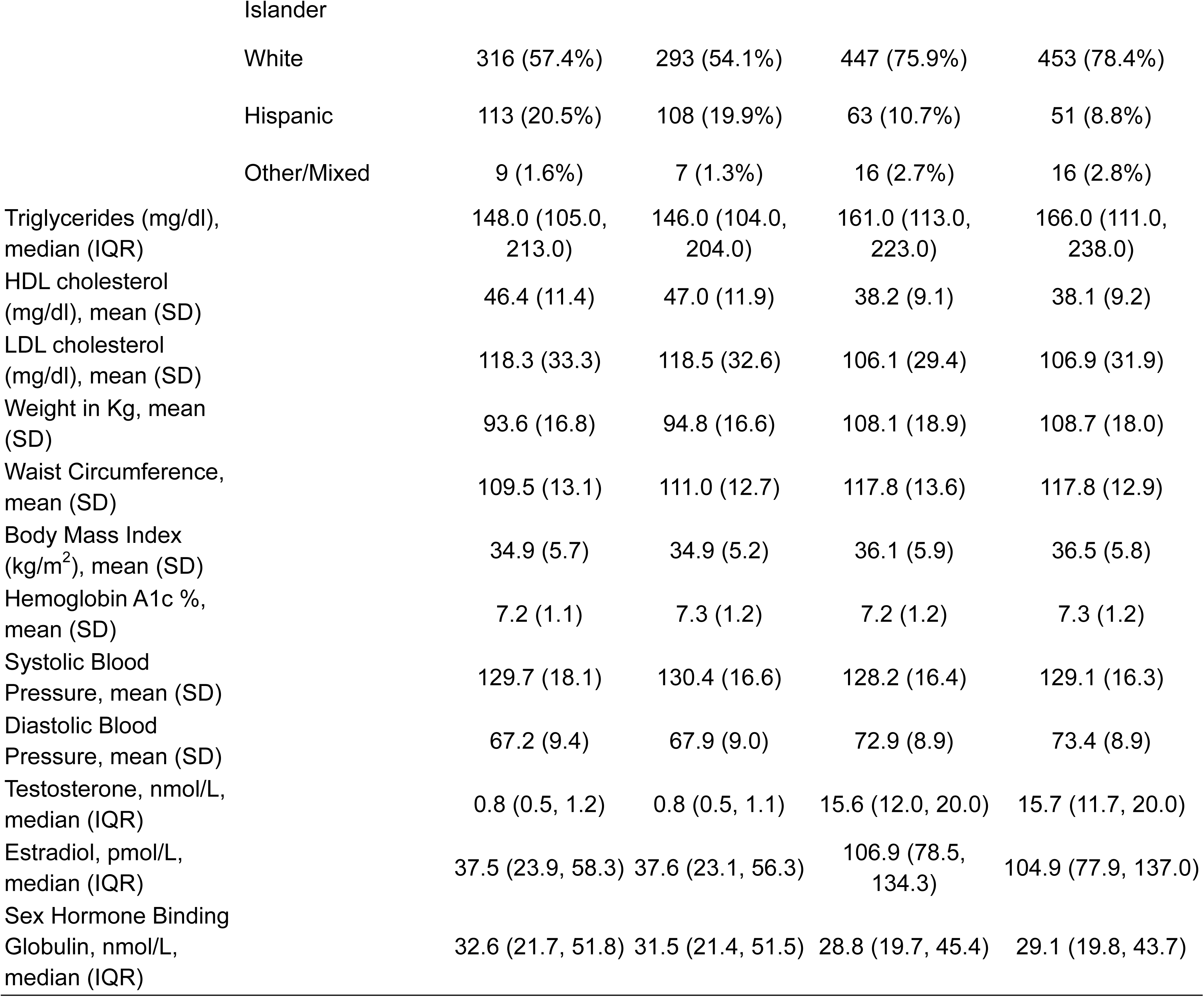
Baseline Characteristics among Postmenopausal Females and Older Males in the Look AHEAD Sex Hormone Ancillary Study (N=2260).

**Supplement Figure 2** shows the percent change in the cardiometabolic risk factors over 8 years of follow up by sex in the ILI and DSE arms. Over 8 years, compared with DSE, the ILI arm achieved significant cardiometabolic improvements in both sexes.^1^

**Supplement Table 1** summarizes the estimated associations between baseline hormone levels and cardiometabolic outcomes across the follow-up years, stratified by randomization arm and sex. For each year we presented the effect of each sex hormone on outcomes in the ILI and DSE treatment groups and the differential effect between ILI vs DSE based on the interaction term (treatment effect).

*Heterogeneity of Treatment Effects (HTE) of Baseline sex Hormones on Cardiometabolic Risk Factors*

### Lipids (TG, LDL-C, HDL-C) (Figure 1, Supplement table 2)

Males with higher baseline E2, and total T experienced TG increases (at year 4, 6 and 8; HTE p = 0.019 for E2, and at year 6 and 8; HTE p = 0.008 for total T) in response to ILI, whereas no similar HTE pattern was observed in females (HTE p =0.800, 0.145, sex difference p = 0.003, 0.001, respectively).

**Figure 1.**
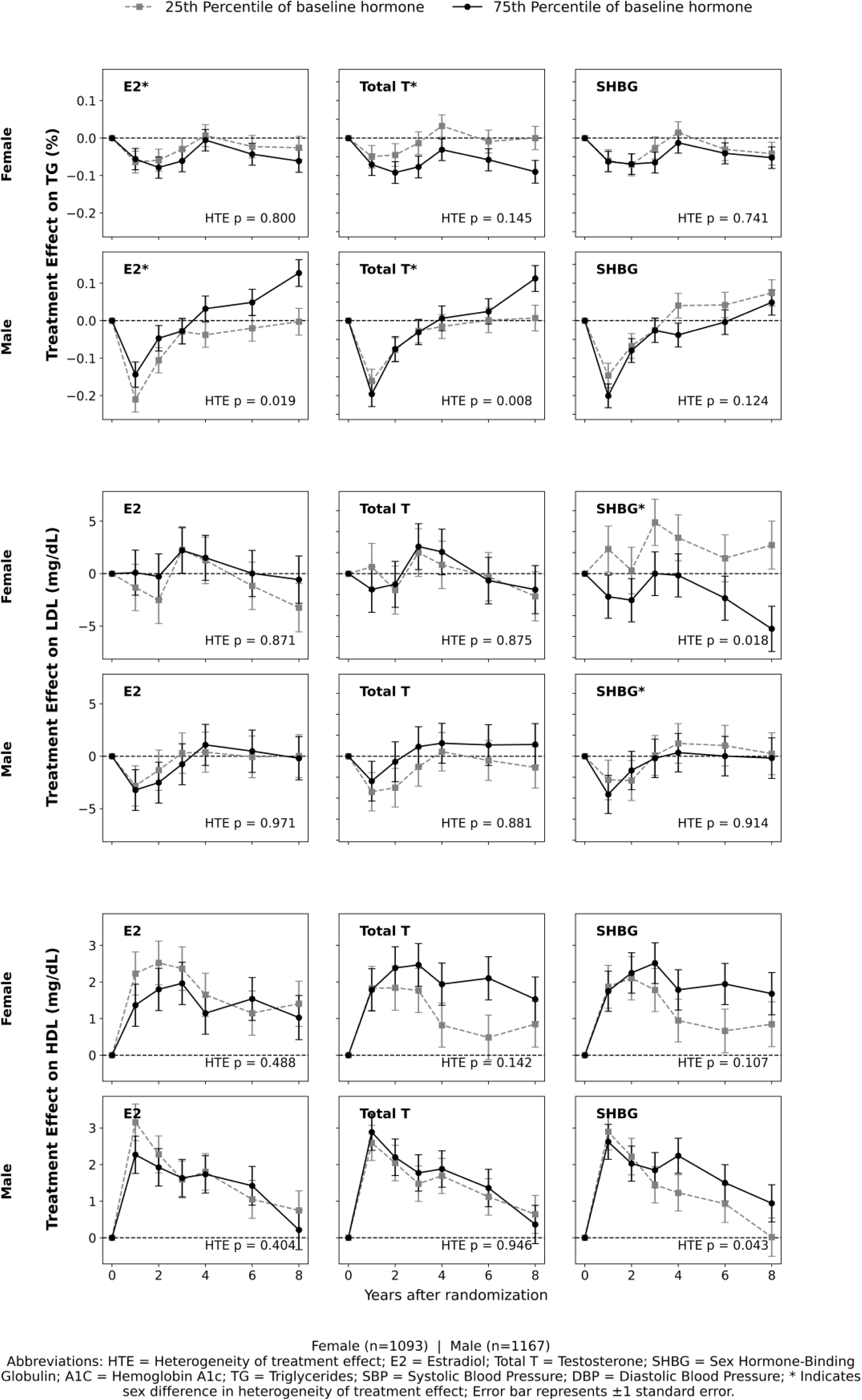
Treatment effects of ILI (vs DSE) on lipids over time.

Females with higher baseline SHBG experienced more LDL-C reduction (HTE p = 0.018) and excluding those with triglycerides ≥400 mg/dL did not change the results (HTE p-value: 0.022), whereas no effect was seen in males (HTE p =0.914, sex difference p = 0.019).

Males with higher baseline SHBG experienced greater HDL-C increases from year 3 onwards (HTE p = 0.043). Similarly, females with higher baseline SHBG experienced greater HDL-C increases beginning in year 3, but it was not statistically significant. (HTE p = 0.107, sex difference p = 0.960)

### Blood Pressure (SBP, DBP) (Figure 2, Supplement table 2)

Females with higher baseline total T experienced greater DBP reduction at year 2 (HTE p=0.030), while a non-statistically significant pattern of treatment effect across varying levels of baseline total T was observed in males (HTE p = 0.181, sex difference p = 0.389). Females with higher baseline SHBG experienced greater SBP reduction, but it was not statistically significant (HTE p=0.054). A different pattern for SBP was seen in males (HTE p = 0.730, sex difference p = 0.001), with no separation of treatment effects on SBP across varying levels of baseline SHBG.

**Figure 2.**
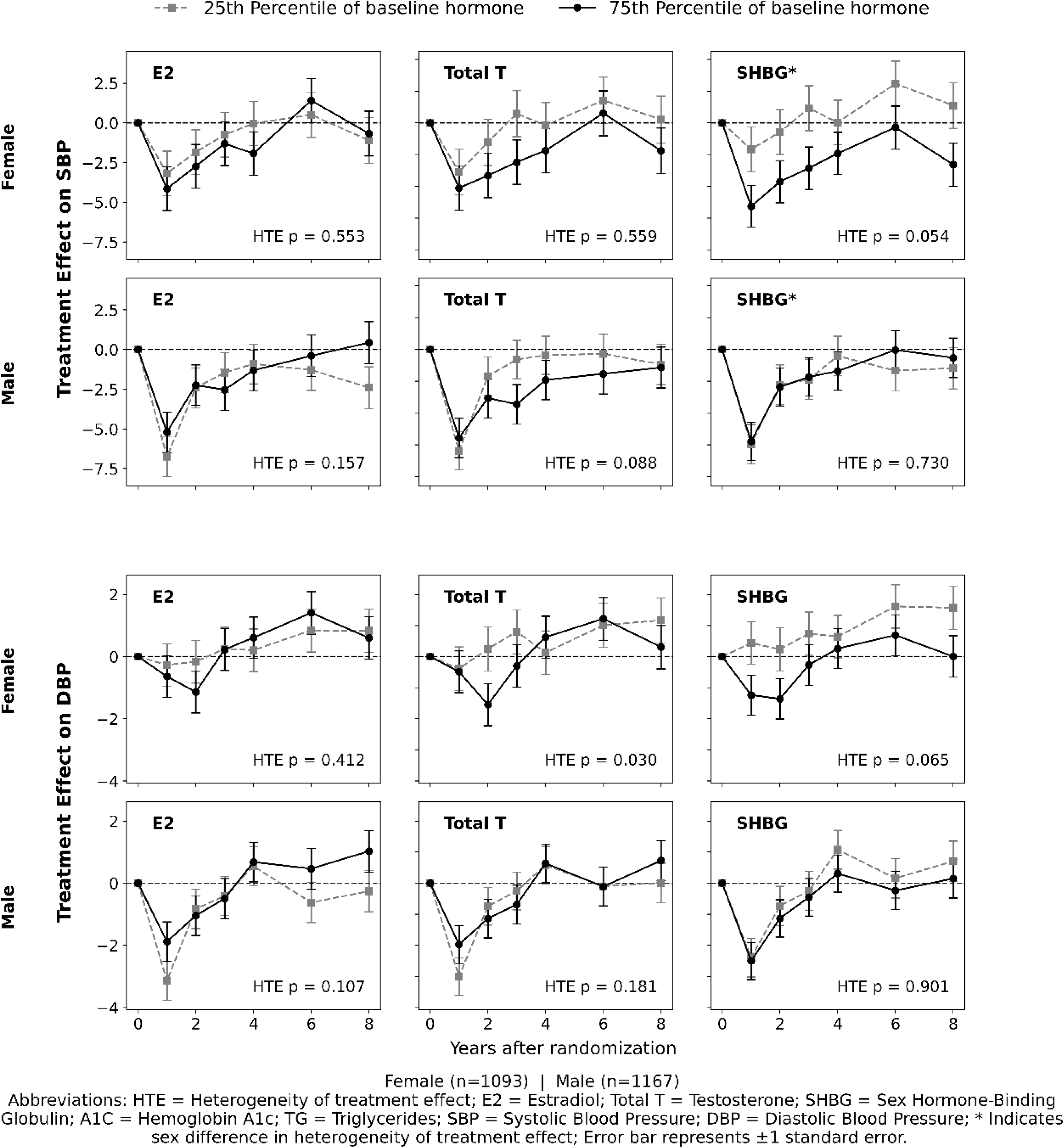
Treatment effects of ILI (vs DSE) on blood pressure over time.

### Body Weight and WC (Figure 3, Supplement table 2)

For weight, both females and males with higher baseline total T experienced greater weight loss, sustained through year 8 in females (HTE p = 0.013) and through year 6 in males (HTE p = 0.003), with interaction by sex (p = 0.049). Males with higher baseline E2 showed greater weight loss through year 6 (HTE p = 0.004). Females showed a similar but non-statistically significant pattern between E2 and weight (HTE p = 0.103, sex difference p = 0.157).

**Figure 3.**
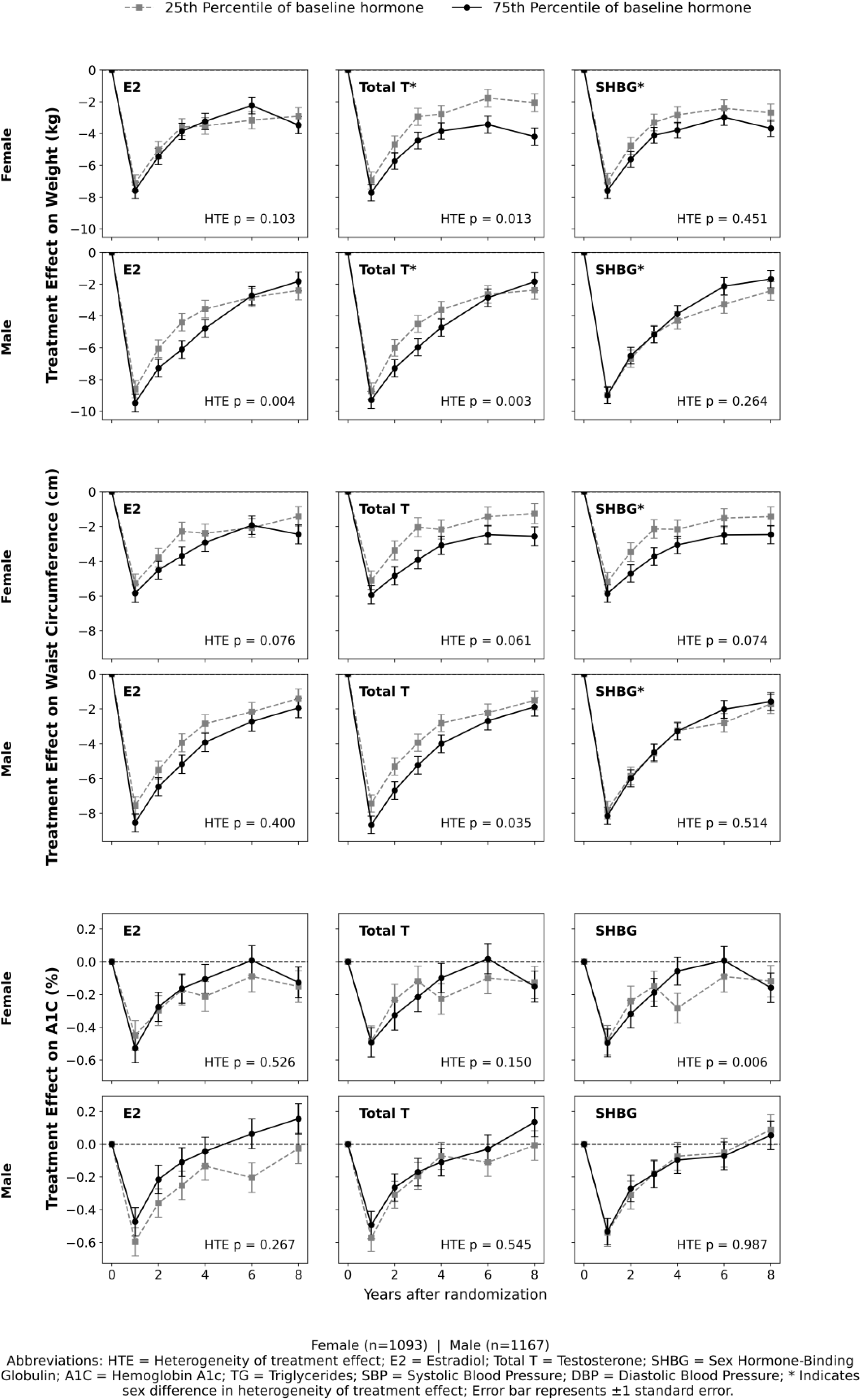
Treatment effects of ILI (vs DSE) on weight, WC, and HbA1c over time.

Higher baseline total T was associated with greater reductions in WC in males (HTE p = 0.035), with similar but non-statistically significant pattern observed for WC in females (HTE p = 0.061, sex difference p = 0.869). SHBG showed a sex difference on effect of ILI on WC (sex difference p= 0.006), with higher SHBG associated with greater WC reduction from year 1 onwards in females (HTE p=0.074). In males, there was no treatment effect across different levels of baseline SHBG (HTE p = 0.514).

### Hemoglobin A1c (HbA1c) (Figure 3, Supplement Table 2)

Females with lower baseline SHBG exhibited steeper HbA1c declines at year 4 and year 6 (HTE p=0.006), with non-statistically significant pattern observed in males (HTE p = 0.987, sex difference p =0.364). Total T mirrored this trend (HTE p=0.150, sex difference p = 0.485). Males with lower E2 demonstrated more HbA1c reduction due to ILI (HTE p=0.267), with similar and non-statistically significant pattern observed in females (HTE p = 0.526, sex difference p= 0.456).

## Discussion

In this secondary analysis of the Look AHEAD trial, we found that the cardiometabolic benefits of an ILI (vs DSE) differed by baseline sex hormone profiles in older adults with T2D, and that these differences were sex specific. Overall, ILI produced significant improvements in weight, and other traditional cardiometabolic risk factors, but the magnitude of changes was modified by baseline sex hormone levels. To our knowledge, this is the first study to assess heterogeneity of treatment effects of an intensive lifestyle intervention by sex hormones and to characterize any sex differences.

### Lipid Outcomes (TG, LDL-C, HDL-C)

Baseline total T modulated the ILI treatment effect on TG in males, while SHBG had effects on LDL-C in females, and HDL-C in males. In females, higher baseline T trended toward steeper ILI-driven TG declines, although not statistically significant (p = 0.145), whereas E2 and SHBG had minimal impact (p>0.7). Conversely, in males, we observed that lower E2 and higher total T at baseline significantly amplified TG reductions during follow-up. These findings are similar to results from cross-sectional studies in cohorts of patients with coronary artery disease. These studies showed that higher circulating E2 correlated with elevated TG in males, and that lower SHBG and lower total T were associated with more atherogenic lipid profiles in males^28–31^.

For unadjusted changes in LDL-C over time, the DSE group benefited slightly more in LDL-C reduction compared to ILI, which was largely attributed to the DSE group’s increased use of lipid-lowering medications, specifically statins^1^. However, in our results, higher baseline SHBG in females was associated with greater LDL-C reduction over time, after statins use adjustment. This finding suggests that higher baseline SHBG levels in females may enhance LDL-C reduction following a lifestyle intervention.

In males, baseline SHBG modulated the HDL-C response to ILI, with higher levels associated with greater HDL-C increases. In females, we observed that higher baseline total T and SHBG were associated with greater HDL-C boosting, similar to prior studies, but the difference did not reach statistical significance^32,33^.

Our finding on the effect of SHBG also supports prior findings of cross-sectional and observational studies that showed SHBG negatively associated with LDL-C and positively associated with HDL-C^34–37^.

### Blood pressure outcomes (SBP, DBP)

Females with higher baseline T and SHBG experienced larger initial reductions in systolic and diastolic blood pressure, but the differences did not reach statistically significance. Benefits in BP reductions were attenuated over the eight years of follow-up. By contrast, male participants’ SBP and DBP trajectories were overlapping across hormone strata, with only a modest sustained SBP reduction linked to higher T (p = 0.088). These sex-specific interactions may reflect the vasodilatory and anti-inflammatory properties of androgens and the role of SHBG as a biomarker of insulin sensitivity and vascular health.

### Weight and WC

Higher baseline T predicted more rapid and durable weight loss due to ILI in both female (p = 0.013) and male participants (p = 0.003). Female participants maintained greater relative reductions in weight through year 8 (sex difference p = 0.049). This supports prior findings in males where higher T was associated with larger weight and metabolic improvements compared to other males with low T^38,39^. Our study demonstrates that postmenopausal females likewise experience superior, longer-lasting weight loss when relatively androgen rich. E2 influenced male weight trajectories. Male participants with higher baseline E2 lost and sustained more weight loss through year 6, reflecting the known positive correlation between adiposity and peripheral E2 production via aromatization in males^40–42^. However, higher E2 may be a marker for greater baseline obesity, capacity for loss rather than directly driving weight change^40^. Higher baseline E2 may support maintenance of weight loss through effects on energy homeostasis and appetite regulation^40,43^.

Both female and male participants with higher baseline total T achieved the largest reductions in central adiposity, consistent with evidence that androgens promote visceral fat mobilization during calorie restriction^44,45^. Other sex hormones had modest WC effects (all HTE p>0.07). The lack of sex difference in total T’s impact on WC suggests that endogenous androgenicity facilitated loss of harmful visceral fat in both males and females. Our results support other studies that showed endogenous testosterone concentrations inversely correlated with central adiposity^46,47^. Modest, non-significant WC effects of E2 and SHBG in our cohort further highlight testosterone as the principal hormonal determinant of ILI-driven central fat reduction.

These results suggest that androgen status at baseline, not sex alone, modulated responsiveness of weight and WC to diet and exercise interventions in T2D.

### Hemoglobin A1C

ILI induced robust HbA1c reductions (∼10% at year 1) that partially rebounded by year 8. Baseline hormone levels did not significantly modify these glycemic benefits in either sex (HTE p > 0.20). This largely uniform response in A1c suggests that weight loss is the driver in glucose control, with sex hormones having a minor role. Our finding that glycemic benefits did not differ by baseline hormone levels corroborates results from smaller trials using testosterone-therapy which similarly showed that glycemic improvements correlate closely with changes in adiposity and muscle mass, rather than initial hormone concentrations^48–53^. From a clinical standpoint, this emphasizes the universal benefit of lifestyle modification for glycemic improvements regardless of sex hormone profiles. It reinforces current guidelines that promote ILI as first-line therapy for glycemic improvements in both males and females with T2D^54,55^.

### Limitations

First, all female participants were postmenopausal; premenopausal or perimenopausal dynamics could differ. Second, other unmeasured hormones like DHEA or progesterone may have related effects which we have not assessed. Third, the ILI was a comprehensive program (diet, exercise, behavioral support); we cannot disentangle which components drove each effect. Finally, medication changes during follow up, though adjusted for, may confound the effects of lipids, A1c and BP.

Our data suggests that pre-intervention sex hormone profiling could help guide the overall approach to recommendations for weight loss in people with T2D. For example, males with lower E2 or higher T may anticipate greater TG-lowering, whereas females with higher SHBG may derive more BP benefits. Incorporating endocrine markers into risk stratification may guide adjunctive lipid lowering treatment, such as targeting TG-lowering in males with higher E2 or antihypertensives in females with lower SHBG. Future research should validate these hormone-intervention interactions in diverse populations, explore underlying mechanisms in lipoprotein and vascular biology, and test combined lifestyle– hormone modulation strategies (e.g., selective androgen receptor modulators, targeted estrogen modulation) to enhance cardiometabolic outcomes.

### Conclusion

Our analysis of Look AHEAD data shows that baseline sex hormones influence ILI benefits in T2D in a sex-specific manner. Males with greater endogenous androgenicity achieved larger reductions in TG, weight, and waist circumference, whereas females with higher total testosterone had greater HDL-C improvements and those with higher SHBG experienced more pronounced blood pressure lowering. These findings extend prior knowledge by identifying hormone-dependent heterogeneity in lifestyle response, and they support a precision medicine approach: assessing a patient’s endocrine status may help personalize lifestyle prescriptions for metabolic health.

## Data availability

Restrictions apply to the availability of some, or all data generated or analyzed during this study to preserve patient confidentiality or because they were used under license. The corresponding author will on request detail the restrictions and any conditions under which access to some data may be provided.

## Funding Sources

This work was funded by NIH-NIDDK grants: R01DK127222 and U01DK57149

The Look AHEAD (Action for Health in Diabetes) trial’s ClinicalTrials.gov number is NCT00017953.

## Acknowledgement

The authors would like to thank Dr. Allen D Everett and the members of his laboratory. We would also thank Dr. David Graham and former members of the Molecular Determinants Core. We also thank all the participants of the Look AHEAD study.

## Nonstandard Abbreviations and Acronyms

T2D: Type 2 Diabetes
ILI: Intensive Lifestyle Intervention
DSE: Diabetes Support and Education
TG: Triglycerides
LDL-C: Low Density Lipoprotein Cholesterol
HDL-C: High Density Lipoprotein Cholesterol
HbA1c: Glycated Hemoglobin
SBP: Systolic Blood Pressure
DBP: Diastolic Blood Pressure
WC: Waist Circumference
Total T: Total Testosterone
E2: Estradiol
SHBG: Sex Hormone–Binding Globulin
GCMS: Gas Chromatography–Mass Spectrometry
ELISA: Enzyme Linked Immunosorbent Assay
HTE: Heterogeneity of Treatment Effect

